# Associations between mask-wearing, handwashing, and social distancing practices and risk of COVID-19 infection in public: a case-control study in Thailand

**DOI:** 10.1101/2020.06.11.20128900

**Authors:** Pawind Doung-ngern, Repeepong Suphanchaimat, Apinya Panjangampatthana, Chawisar Janekrongtham, Duangrat Ruampoom, Nawaporn Daochaeng, Napatchakorn Eungkanit, Nichakul Pisitpayat, Nuengruethai Srisong, Oiythip Yasopa, Patchanee Plernprom, Pitiphon Promduangsi, Panita Kumphon, Paphanij Suangtho, Peeriya Watakulsin, Sarinya Chaiya, Somkid Kripattanapong, Thanawadee Chantian, Emily Bloss, Chawetsan Namwat, Direk Limmathurotsakul

**Author notes:** **Corresponding author:** Direk Limmathurotsakul, 420/6 Mahidol-Oxford Tropical Medicine Research Unit, Faculty of Tropical Medicine, Rajvithi Road, Bangkok, Thailand, 10400. **Alternative corresponding author:** Pawind Doungngern, Department of Disease Control, Ministry of Public Health, Nonthaburi, 11000, Thailand. **Disclaimer:** The findings and conclusions in this report are those of the authors and do not necessarily represent the official position of the Centers for Disease Control and Prevention.

## Abstract

We evaluated the effectiveness of personal protective measures, including mask-wearing, handwashing, and social distancing, against COVID-19 infection among contacts of cases. We conducted a case-control study with 211 cases and 839 non-matched controls using all contact tracing records of Thailand’s national Surveillance and Rapid Response Team. Cases were asymptomatic contacts of COVID-19 patients identified between 1 and 31 March 2020 who were diagnosed with COVID-19 by 21 April 2020; controls were asymptomatic contacts who were not diagnosed with COVID-19. Participants were queried about practices during contact periods with a case. Adjusted odds ratios (aOR) and 95% confidence intervals (CI) were estimated for associations between diagnosis of COVID-19 and covariates using multivariable logistic regression models. Wearing masks all the time during contact was independently associated with lower risk of COVID-19 infection compared to not wearing masks (aOR 0.23, 95% CI 0.09– 0.60), while sometimes wearing masks during contact was not (aOR 0.87, 95% CI 0.41–1.84). Maintaining at least 1 meter distance from a COVID patient (aOR 0.15, 95% CI 0.04–0.63), duration of close contact ≤15 minutes versus longer (aOR 0.24, 95% CI 0.07–0.90), and handwashing often (aOR 0.34, 95% CI 0.13–0.87) were significantly associated with lower risk of infection. Type of mask was not independently associated with infection. Those who wore masks all the time also were more likely to practice social distancing. Our findings suggest consistent wearing of masks, handwashing, and social distancing in public to protect against COVID-19 infection.

## Introduction

Evaluation of the effectiveness of mask-wearing by healthy persons in the general public against COVID-19 infection is urgently needed (*1, 2*). On 27 February 2020, during the early stages of the COVID-19 outbreak, the World Health Organization (WHO) announced “For asymptomatic individuals, wearing a mask of any type is not recommended” (*3*). The rationale, at that time, was to avoid unnecessary cost, procurement burden, and a false sense of security (*3*). A number of systematic reviews found no conclusive evidence to support widespread use of masks in public against respiratory infectious diseases, such as influenza and SARS (*4-6*). However, China and many countries in Asia including South Korea, Japan, and Thailand have recommended the use of face masks among the general public since early in the outbreak (*7*). There is evidence that COVID-19 patients can have a “pre-symptomatic” period, during which infected persons can be contagious and, therefore, transmit the virus to others before symptoms develop (*8*). These findings led to a change in recommendations from the US Centers for Disease Control and Prevention on 4 April 2020 that advised all persons to wear a cloth face covering when in public (*9*). On 6 April and 5 June 2020, WHO updated its advice on the use of masks for the general public, and encouraged countries that issue the recommendations to conduct research on this topic (*8*).

Thailand has been implementing multiple measures against transmission of COVID-19 since the beginning of the outbreak (*10, 11*). The country established thermal screening at airports on 3 January 2020, and detected the first case of COVID-19 outside China—a traveler from Wuhan arriving at Bangkok Suvarnabhumi airport—on 8 January 2020 (*10*). The country utilizes Surveillance and Rapid Response Teams (SRRTs), together with village health volunteers, to conduct contact tracing, educate the public about the disease, and monitor the close contacts of COVID-19 patients in quarantine (*11*). SRRTs are epidemiologic investigation teams trained to conduct surveillance, investigations, and initial controls of communicable diseases including influenza H5N1, severe acute respiratory syndrome (SARS), and Middle East respiratory syndrome (*12, 13*). Currently, there are more than 1,000 SRRTs established at district, provincial, and regional levels in Thailand (*12*) working on COVID-19 contact tracing.

By February 2020, public pressure to wear masks in Thailand was high, medical masks had become difficult for the public to procure, and the government had categorized medical masks as price-controlled goods and announced COVID-19 as a dangerous communicable disease according to the Communicable Disease Act 2015. These developments empowered officials to quarantine contacts and close public venues (*14, 15*). On 3 March, the Thai Ministry of Public Health (MoPH) announced that public use of cloth face masks were recommended (*16*). On 18 March, schools, universities, bars, nightclubs and entertainment venues were closed (*17*). On 26 March, when the country was reporting ∼100–150 new COVID-19 patients per day, the government declared a national state of emergency, prohibited public gatherings, and enforced wearing of face masks by all persons on public transport (*18*). On 21 April, 19 new PCR-confirmed COVID-19 patients were announced by MoPH, bringing the total number of patients to 2,811 (*18*).

Given the current lack of evidence, we sought to evaluate the effectiveness of mask wearing, handwashing, social distancing, and other preventive measures against COVID-19 infection in Thailand.

## Methods

### Study design

We conducted a retrospective case-control study in which both cases and controls were drawn from a cohort of contact tracing records of the central SRRT team at the Department of Disease Control (DDC), MoPH, Thailand (Figure 1). Contacts were defined by the DDC MoPH as individuals who had activities together with or were in the same location(s) as a COVID-19 patient (*19, 20*). The main aim of contact tracing was to identify and evaluate contacts, perform reverse transcription polymerase chain reaction (RT-PCR) COVID-19 diagnostic tests, and quarantine high-risk contacts, as defined by the MoPH (Table 1). Contacts were classified as high-risk if they were family members or lived in the same household as a COVID-19 patient, if they were within a 1-meter distance of a COVID-19 patient longer than 15 minutes; if they were exposed to coughs, sneezes, or secretions of a COVID-19 patient and were not wearing protective gear, such as a mask; or if they were in the same closed environment within a 1-meter distance of a COVID-19 patient longer than 15 minutes and were not wearing protective gear, such as a mask (*19, 20*). All RT-PCR tests were performed at laboratories certified for COVID-19 testing by the National Institute of Health of Thailand (*19, 20*). Data on risk factors associated with COVID-19 infection, such as type of contact and use of mask, were recorded during the contact investigation, but data were sometimes incomplete.

**Table 1.**
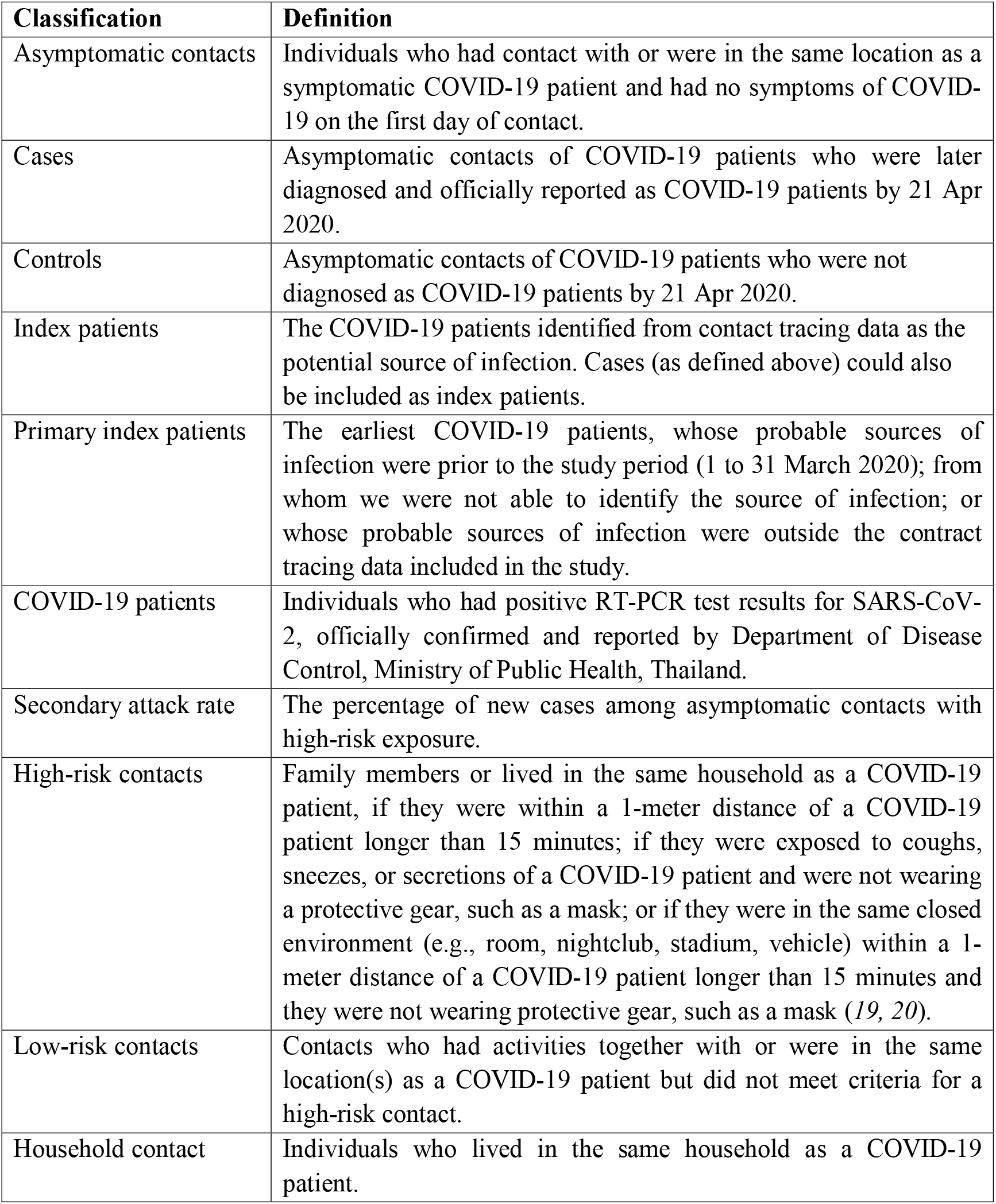
Definitions used in the study.

| **Classification** | **Definition** |
| --- | --- |
| Asymptomatic contacts | Individuals who had contact with or were in the same location as a symptomatic COVID-19 patient and had no symptoms of COVID-19 on the first day of contact. |
| Cases | Asymptomatic contacts of COVID-19 patients who were later diagnosed and officially reported as COVID-19 patients by 21 Apr 2020. |
| Controls | Asymptomatic contacts of COVID-19 patients who were not diagnosed as COVID-19 patients by 21 Apr 2020. |
| Index patients | The COVID-19 patients identified from contact tracing data as the potential source of infection. Cases (as defined above) could also be included as index patients. |
| Primary index patients | The earliest COVID-19 patients, whose probable sources of infection were prior to the study period (1 to 31 March 2020); from whom we were not able to identify the source of infection; or whose probable sources of infection were outside the contact tracing data included in the study. |
| COVID-19 patients | Individuals who had positive RT-PCR test results for SARS-CoV-2, officially confirmed and reported by Department of Disease Control, Ministry of Public Health, Thailand. |
| Secondary attack rate | The percentage of new cases among asymptomatic contacts with high-risk exposure. |
| High-risk contacts | Family members or lived in the same household as a COVID-19 patient, if they were within a 1-meter distance of a COVID-19 patient longer than 15 minutes; if they were exposed to coughs, sneezes, or secretions of a COVID-19 patient and were not wearing a protective gear, such as a mask; or if they were in the same closed environment (e.g., room, nightclub, stadium, vehicle) within a 1-meter distance of a COVID-19 patient longer than 15 minutes and they were not wearing protective gear, such as a mask (19, 20). |
| Low-risk contacts | Contacts who had activities together with or were in the same location(s) as a COVID-19 patient but did not meet criteria for a high-risk contact. |
| Household contact | Individuals who lived in the same household as a COVID-19 patient. |

**Figure 1.**
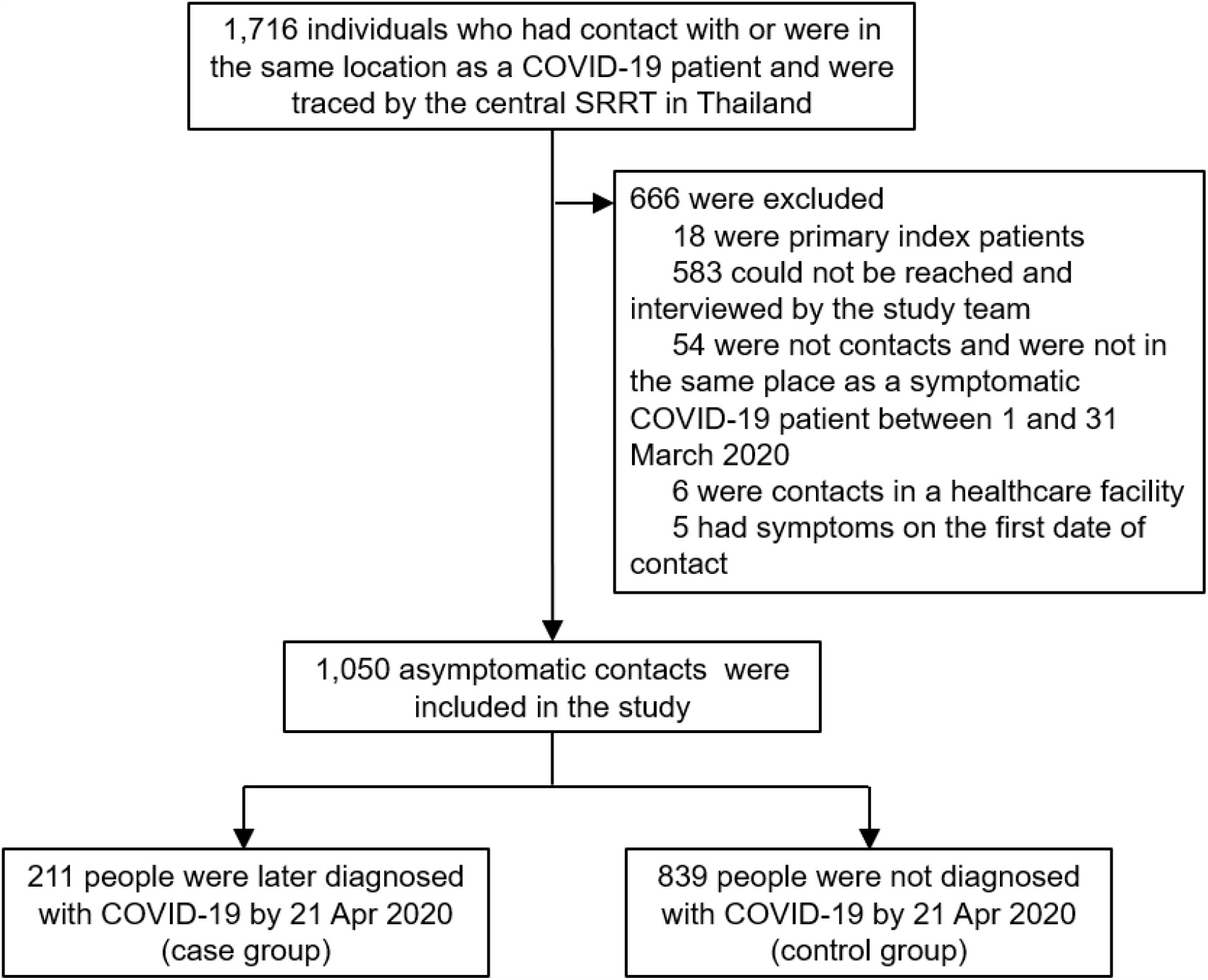
Study flow diagram. **Footnote to Figure 1**. SRRT= Surveillance and Rapid Response Team (SRRT), Ministry of Public Health (MoPH), Thailand

The central SRRT team performed contact investigations for any cluster with at least five PCR-confirmed COVID-19 patients from the same location(s) within a 1-week period (*19, 20*). We used these data to identify contacts of COVID-19 patients who were asymptomatic from 1 through 31 March 2020. All available contact tracing records of the central SRRT team were used in the study. See Supplemental Materials for further details.

We telephoned the contacts during 30 April–27 May 2020 and asked details about their contact with COVID-19 patients (e.g., date(s), location(s), duration and distance of contacts); whether, during the contact period, they wore a mask and the type of mask (1) never, 2) yes, non-medical mask, 3) yes, medical mask, 4), yes, alternately non-medical and medical masks, 5) unknown or cannot remember); and if so, the frequency of wearing a mask during the contact period (1) sometimes, 2) all the time, 3) unknown or cannot remember); if and how frequently they washed their hands during the contact period (1) no washing with soap or alcohol-based solutions, 2) yes, sometimes, 3) yes, all the time after any contact (defined below as ‘often’), 4) unknown or cannot remember); if they performed social distancing, including type of contact with COVID-19 patient or other people at place of contact, if unable to remember who the patient was (1) had physical contact, 2) shortest distance ≤1 meter and no physical contact, 3) shortest distance >1 meter and no physical contact, 4) unknown or cannot remember); total duration of contact (1) in total, more than 1 hour, 2) at least 15 minutes but not more than 1 hour, 3) less than 15 minutes, 4) unknown or cannot remember); if they shared a cup or a cigarette with other people in the place they had contact or had highest risk of contact with the patient (1) no, 2) yes, 3) unknown or cannot remember); and whether the COVID-19 patient, if known to the respondent, had worn a mask (1) never, 2) yes, non-medical mask, 3) yes, medical mask, 4) yes, alternately non-medical and medical masks, 5) unknown or cannot remember), and if so, the frequency of wearing a mask (1) sometimes, 2) all the time, 3) unknown or cannot remember). We also asked, and verified using DDC records, whether and when the contacts became sick and diagnosed with COVID-19.

We defined cases as asymptomatic contacts who were later diagnosed with COVID-19 based on RT-PCR assay results available by 21 Apr 2020 (Table 1). All asymptomatic contacts who were not diagnosed with COVID-19 by 21 Apr 2020, including those who tested negative and those who were not tested, were classified as controls. We used 21 days after 31 March as a cutoff date based on evidence that (1) most COVID-19 patients would likely develop symptoms within 14 days (*21*) and (2) it could take <7 additional days for symptomatic patients under contact investigation to present at healthcare facilities and be tested for COVID-19 by RT-PCR assay.

Definitions for asymptomatic contacts, cases, controls, index patients, primary index patients, COVID-19 patients, high-risk and low-risk contacts were described in Table 1. The reporting of this study follows the STROBE guidelines (see Supplemental Materials) (*22*).

### Statistical analysis

To include only initially asymptomatic contacts in the study, we excluded from the analysis people who reported having any symptoms of COVID-19 at the time of initial contact with a case. We excluded contacts whose contact locations were healthcare facilities because this study focused on the risk of infection in the community. Primary index patients were also excluded if they were the first to have symptoms at the contact investigation location, had symptoms since the first day of visiting the location, or were the origin of infection based on the contact investigation.

Univariate and bivariate analyses were conducted. We estimated secondary attack rate using definitions as described in Table 1 to allow for comparison with other studies. Odds ratios (OR) and 95% confidence intervals (CI) were estimated for associations between development of COVID-19 and baseline covariates, such as mask-wearing, handwashing, and social distancing during the contact period. We used logistic regression with random effects for location and for index patient nested within the same location. If an asymptomatic contact had contact with ≥1 symptomatic COVID-19 patient, the interviewer identified the index patient as the symptomatic COVID-19 patient who had the closest contact. The percentage of missing values for the variable indicating whether the index patients wore a mask was 27%, so this variable was not included in analyses. For other variables, we assumed that missing values were missing at random and used imputation by chained equations (*23, 24*). We created 10 imputed datasets and the imputation model included the case-control indicator and all independent variables included in the multivariable models. We developed the final multilevel mixed-effect logistic regression models on the basis of previous knowledge and a purposeful selection method (*25*). Because of collinearity between mask use and mask type, we conducted a separate analysis including mask type in the multilevel mixed-effects logistic regression model for COVID-19 infection. We also tested a pre-defined interaction between mask type and mask-wearing compliance (see Supplemental Materials).

To better understand patterns of behavior and factors related to compliance in mask-wearing, we also estimated OR and 95% CI for associations between three categories of mask-wearing compliance (i.e. never, sometimes, all the time) and other practices including handwashing and social distancing during the contact period using multinomial logistic regression models with the imputed data set. Logistic regression was used to estimate a p-value for pairwise comparisons.

To estimate the proportional reduction in cases that would occur if exposure to risk factors was reduced, we estimated the population attributable fraction (PAF) using the imputed dataset and a direct method based on logistic regression, as described previously (see Supplemental Materials) (*26, 27*). The final multivariable model was modified by considering each risk factor dichotomously, and PAF was calculated by subtraction of the total number of predicted cases from total number of observed cases, divided by the total number of observed cases.

STATA version 14.2 and R version 4.0.0 were used for all analyses.

## RESULTS

### Characteristics of the cohort data

The contact tracing records of the central SRRT team included 1,716 persons who had contact with or were in the same location as a COVID-19 patient who was diagnosed in an investigation of three large clusters in nightclubs, boxing stadiums, and a state enterprise office in Thailand (Figure 1). Overall, 18 individuals were defined as primary index patients: 11 from the nightclub cluster, 5 from the boxing stadium cluster, and 2 from the state enterprise office cluster. Timelines of primary index patients from nightclub, boxing stadium, and state enterprise clusters are described in detail in the Supplemental Text and Supplemental Figures 1–3. All 18 primary index patients were excluded from case-control study analyses.

### Characteristics of cases and controls

After retrospectively interviewing each contact by phone and applying exclusion criteria (Figure 1), we included 1,050 asymptomatic contacts who had contact with or were in the same location as a symptomatic COVID-19 patient from 1 through 31 March 2020 in the analysis. The median age of individuals was 38 years (interquartile range 28–51) and 55% were male (Table 2). Most asymptomatic contacts included in the study were associated with the boxing stadium cluster (61%, n=645); 36% (n=374) were related to the nightclub cluster, and 3% (n=31) were related to the state enterprise office cluster. Overall, 890 (85%) were contacts with high-risk exposure.

**Table 2.** Factors associated with COVID-19 infection among persons followed through contract tracing, Thailand, March-April 2020.

| **Factors** | **Cases**<br>**(n=211)** | **Controls**<br>**(n=839)** | **Crude odds ratio**<br>**(95% CI)<sup>a</sup>** | **P** | **Adjusted odds**<br>**ratio (95% CI)<sup>a</sup>** | **P** |
| --- | --- | --- | --- | --- | --- | --- |
| **Male gender** | 146/211 (69%) | 434/838 (52%) | 0.83 (0.47–1.46) | 0.52 | 0.76 (0.41–1.41) | 0.37 |
| **Age group** |  |  |  |  |  |  |
| ≤15 years old | 6/211 (3%) | 49/829 (6%) | 0.65 (0.17–2.48) | 0.29 | 0.57 (0.15–2.21) | 0.21 |
| >15–40 years old | 94/211 (45%) | 435/829 (52%) | 1.0 |  | 1.0 |  |
| >40–65 years old | 98/211 (46%) | 302/829 (36%) | 1.65 (0.91–2.97) |  | 1.77 (0.94–3.32) |  |
| >65 years old | 13/211 (6%) | 43/829 (5%) | 1.29 (0.33–5.07) |  | 0.97 (0.22–4.24) |  |
| **Contact place<sup>b</sup>** |  |  |  |  |  |  |
| Nightclub | 35 (17%) | 193 (23%) | Not applicable<sup>c</sup> | - | Not applicable<sup>c</sup> | - |
| Boxing stadium | 125 (59%) | 19 (2%) |  |  |  |  |
| Workplace | 11 (5%) | 286 (34%) |  |  |  |  |
| Household | 38 (18%) | 192 (23%) |  |  |  |  |
| Others | 2 (1%) | 149 (18%) |  |  |  |  |
| **Shortest distance of contact** |  |  |  |  |  |  |
| Physical contact | 132/197 (67%) | 292/809 (36%) | 1.0 | 0.001 | 1.0 | 0.02 |
| ≤1 meter without physical contact | 61/197 (31%) | 335/809 (41%) | 0.76 (0.43–1.35) |  | 1.09 (0.58–2.07) |  |
| >1 meter | 4/197 (2%) | 182/809 (22%) | 0.08 (0.02–0.31) |  | 0.15 (0.04–0.63) |  |
| **Duration of contact within 1 meter** |  |  |  |  |  |  |
| >60 minutes | 180/199 (90%) | 487/801 (61%) | 1.0 | 0.003 | 1.0 | 0.09 |
| >15–60 minutes | 14/199 (7%) | 162/801 (20%) | 0.53 (0.24–1.17) |  | 0.67 (0.29–1.55) |  |
| ≤15 minutes | 5/199 (3%) | 152/801 (19%) | 0.13 (0.04–0.46) |  | 0.24 (0.07–0.90) |  |
| **Sharing dishes or cups<sup>d,e</sup>** |  |  |  |  |  |  |
| None | 125/210 (60%) | 576/837 (69%) | 1.0 | 0.001 | 1.0 | 0.39 |
| Yes | 85/210 (40%) | 261/837 (31%) | 2.71 (1.48–4.94) |  | 1.33 (0.70–2.54) |  |
| **Sharing cigarettes<sup>d,f</sup>** |  |  |  |  |  |  |
| None | 196/209 (94%) | 824/836 (99%) | 1.0 | 0.001 | 1.0 | 0.03 |
| Yes | 13/209 (6%) | 12/836 (1%) | 6.12 (2.12–17.72) |  | 3.47 (1.09–11.02) |  |
| **Washing hands<sup>d,g</sup>** |  |  |  |  |  |  |
| None | 44/210 (21%) | 121/826 (15%) | 1.0 | <0.001 | 1.0 | 0.045 |
| Sometimes | 114/210 (54%) | 333/826 (40%) | 0.41 (0.18–0.91) |  | 0.34 (0.14–0.81) |  |
| Often | 52/210 (25%) | 372/826 (45%) | 0.19 (0.08–0.46) |  | 0.33 (0.13–0.87) |  |
| **Wearing masks<sup>d,h</sup>** |  |  |  |  |  |  |
| Not wearing masks | 102/211 (48%) | 500/834 (60%) | 1.0 | 0.003 | - | - |
| Wearing non-medical masks | 25/211 (12%) | 77/834 (9%) | 0.78 (0.32–1.90) |  |  |  |
| Wearing non-medical and medical masks alternately | 12/211 (6%) | 48/834 (6%) | 0.46 (0.13–1.64) |  |  |  |
| Wearing medical masks | 72/211 (34%) | 209/834 (25%) | 0.25 (0.12–0.53) |  |  |  |
| **Compliance with mask wearing<sup>d,h</sup>** |  |  |  |  |  |  |
| Not wearing a mask | 102/210 (49%) | 500/823 (61%) | 1.0 | <0.001 | 1.0 | 0.006 |
| Sometimes | 79/210 (38%) | 125/823 (15%) | 0.75 (0.37–1.52) |  | 0.87 (0.41–1.84) |  |
| All the time | 29/210 (14%) | 198/823 (24%) | 0.16 (0.07–0.36) |  | 0.23 (0.09–0.60) |  |

<sup>a</sup> Both crude and adjusted odds ratios were estimated using logistic regression with a random effect for location and a random effect for index patient nested within the same location. <sup>b</sup> The state enterprise office was included as a workplace. “Others” included restaurants, markets, malls, religious places, and households of index patients or other people but not living together. <sup>c</sup> Location was included in the model as a random effect variable. <sup>d</sup> During the contact period. <sup>e</sup> Sharing multi-serving dishes using communal serving utensils to portion food individually was categorized as not sharing dishes. <sup>f</sup> Included sharing electronic cigarettes and any vaping devices. <sup>g</sup> Included washing with soap and water, and with alcohol-based solutions. <sup>h</sup> Wearing masks incorrectly (e.g., not covering both nose and mouth) was considered to be the same as not wearing a mask for analyses.

In total, 211 (20%) asymptomatic contacts were later diagnosed with COVID-19 by 21 Apr 2020 and classified as cases; 839 (80%) were not diagnosed and represented the control group. Of the 211 cases, 195 (92%) were contacts with high-risk exposure and 150 (71%) had symptoms of COVID-19 prior to diagnosis by RT-PCR results. The last date that a COVID-19 case was diagnosed was 9 April 2020. Among 839 controls, 696 (83%) were deemed to have had high-risk exposures and 719 (86%) were tested for COVID-19 at least once.

Figure 2 illustrates contacts (and possible transmission routes) between index patients and asymptomatic contacts included in the study. There were 228, 144, and 20 asymptomatic contacts of index patients at nightclubs, boxing stadiums, and the state enterprise office, respectively. Figure 2 shows all contacts of primary index patients in the clusters. The others had contact with cases associated with nightclubs, boxing stadiums, at workplaces (n=277), households (n=230), and other places (n=151).

**Figure 2.**
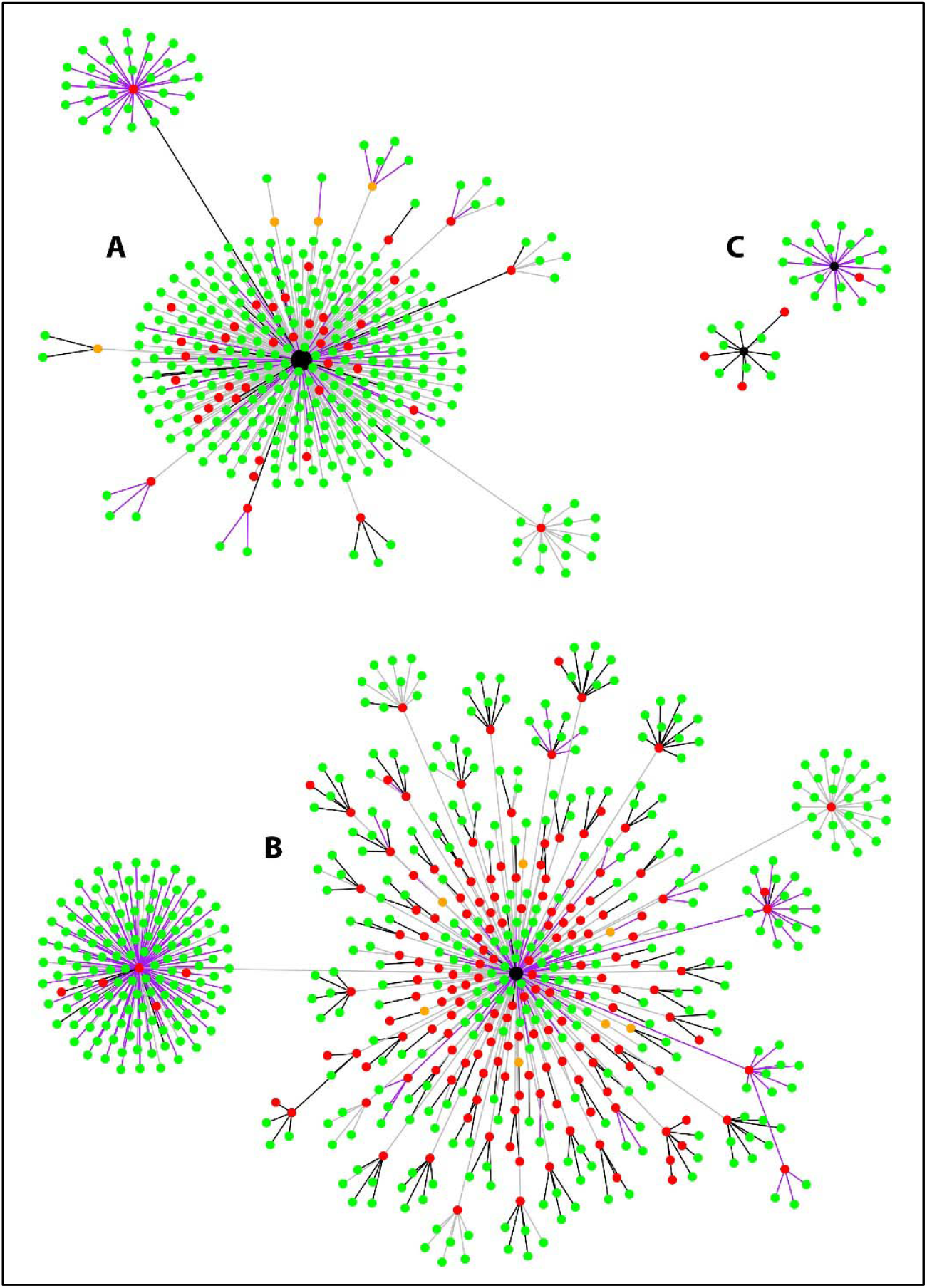
Development and transmission of COVID-19 among asymptomatic contacts in Thailand, during March-April 2020, included in the study. **Footnote to Figure 2**. A, B, and C represent the nightclub cluster, boxing stadium cluster, and state enterprise office cluster, respectively. Black nodes represent primary index patients, red dots represent cases, and green dots represent controls. Orange dots represent index patients (confirmed COVID-19 patients) who could not be contacted by the study team. Black lines represent household contacts, purple lines represent contacts at workplaces, and gray lines represent contacts at other locations. Definitions of index patients, cases, and controls are listed in Table 1.

Among 890 asymptomatic contacts with high-risk exposures included in the study, the boxing stadium secondary attack rate was 86% (111/129), the nightclub secondary attack rate was 18% (34/187), the household secondary attack rate was 17% (38/230), the workplace secondary attack rate was 5% (10/205), and the secondary attack rate at other places was 1% (2/139).

### Bivariate analyses

Table 2 shows that COVID-19 infection was negatively associated with maintaining a distance of at least >1 meter from a case (crude odds ratio [OR] 0.08, 95% CI 0.02–0.31), duration of contact ≤15 minutes (OR 0.13, 95% CI 0.04–0.46), handwashing sometimes (OR 0.41, 95% CI 0.18–0.91) or often (OR 0.19, 95% CI 0.08–0.46) and wearing a mask all the time during the period of contact with the COVID-19 case (OR 0.16, 95% CI 0.07–0.36). Sharing dishes or cups (OR 2.71 95%CI 1.48–4.94) and sharing cigarettes (OR 6.12, 2.12–17.72) with other people (not necessarily including the case) were associated with higher risk of COVID-19 infection. Type of mask was associated with infection in the bivariate model (p=0.003).

### Multivariable analyses

We found a negative association between risk of COVID-19 infection and maintaining a distance of at least >1 meter from a case (adjusted odds ratio [aOR] 0.15, 95% CI 0.04–0.63), duration of contact ≤15 minutes (aOR 0.24, 95% CI 0.07–0.90), handwashing often (aOR 0.34, 95% CI 0.13–0.87), and wearing a mask all the time during the period of contact with the COVID-19 case (aOR 0.23, 95% CI 0.09–0.60) (Table 2). Wearing masks sometimes during contact with the case was not significantly associated with lower risk of infection (aOR 0.87, 95% CI 0.41–1.84). Sharing cigarettes with other people was associated with higher risk of COVID-19 infection (aOR 3.47, 1.09–11.02).

Compliance with mask-wearing during the period of contact with a case was strongly associated with lower risk of infection in the multivariable model. Because of collinearity with mask-wearing compliance, mask type was not included in the final model. We included mask type in a separate multivariable model and found type of mask was not independently associated with infection (p=0.54) (Supplemental Materials, Table 1). We found no evidence of effect modification between mask type and mask-wearing compliance.

### Association between mask-wearing compliance and other social distancing practices

Because mask-wearing throughout the contact period was negatively associated with COVID-19 infection, we further explored characteristics of those patients to ascertain if wearing a mask produced a potential false sense of security. We found that during the contact period, those who wore masks all the time were more likely to report closest contacts >1 meter (25% vs. 18%, pairwise p=0.03), durations of contact ≤15 minutes (26% vs 13%, pairwise p<0.001), and washing hands often (79% vs. 26%, pairwise p<0.001) compared with those who did not wear masks (Table 3). We found that those who wore masks sometimes were more likely to wash their hands often (43% vs. 26%, pairwise p<0.001) compared with those who did not wear masks. However, they were more likely to have physical contact (50% vs. 42%, pairwise p=0.03) and report duration of contacts >60 minutes (75% vs. 67%, pairwise p=0.04) compared with those who did not wear masks.

**Table 3.** Factors associated with mask-wearing compliance among persons followed through contract tracing, Thailand, March-April 2020.

| Factors | Not wearing masks<br>(n=602) | Wearing masks sometimes<br>(n=204) | Wearing masks all the time<br>(n=227) | P |
| --- | --- | --- | --- | --- |
| **Male gender** | 324/601 (54%) | 129/204 (63%) | 115/227 (51%) | 0.03 |
| **Age group** |  |  |  |  |
| ≤15 years old | 45/594 (8%) | 5/204 (2%) | 3/225 (1%) | <0.001 |
| >15–40 years old | 269/594 (45%) | 117/204 (57%) | 132/225 (59%) |  |
| >40–65 years old | 236/594 (40%) | 76/204 (37%) | 84/225 (37%) |  |
| >65 years old | 44/594 (7%) | 6/204 (3%) | 6/225 (3%) |  |
| **Contact places** |  |  |  |  |
| Nightclub | 84 (14%) | 51 (25%) | 91 (40%) | <0.001 |
| Boxing stadium | 48 (8%) | 66 (32%) | 29 (13%) |  |
| Workplace <sup>a</sup> | 178 (30%) | 46 (23%) | 64 (28%) |  |
| Household | 167 (28%) | 27 (13%) | 33 (15%) |  |
| Others <sup>b</sup> | 125 (21%) | 14 (7%) | 10 (4%) |  |
| **Shortest distance of contact** |  |  |  |  |
| Physical contact | 246/588 (42%) | 96/191 (50%) | 76/212 (36%) | 0.005 |
| ≤1 meter without physical contact | 238/588 (40%) | 70/191 (37%) | 83/212 (39%) |  |
| >1 meter | 104/588 (18%) | 25/191 (13%) | 53/212 (25%) |  |
| **Duration of contact within 1 meter** |  |  |  |  |
| >60 minutes | 396/590 (67%) | 143/190 (75%) | 121/205 (59%) | <0.001 |
| >15–60 minutes | 120/590 (20%) | 23/190 (12%) | 30/205 (15%) |  |
| ≤15 minutes | 74/590 (13%) | 24/190 (13%) | 54/205 (26%) |  |
| **Sharing dishes or cups <sup>c,d</sup>** |  |  |  |  |
| None | 361/601 (60%) | 130/203 (64%) | 200/226 (88%) | <0.001 |
| Yes | 240/601 (40%) | 73/203 (36%) | 26/226 (12%) |  |
| **Sharing cigarettes <sup>c,e</sup>** |  |  |  |  |
| None | 586/600 (98%) | 194/202 (96%) | 223/226 (99%) | 0.29 |
| Yes | 14/600 (2%) | 8/202 (4%) | 3/226 (1%) |  |
| **Washing hands <sup>c,f</sup>** |  |  |  |  |
| None | 142/594 (24%) | 16/203 (8%) | 6/224 (3%) | <0.001 |
| Sometimes | 298/594 (50%) | 99/203 (49%) | 42/224 (19%) |  |
| Often | 154/594 (26%) | 88/203 (43%) | 176/224 (79%) |  |

P values were estimated using univariable multinomial logistic regression models. Missing values were imputed using the imputation model. Wearing masks incorrectly (e.g., not covering both nose and mouth) was considered to be the same as not wearing a mask for analyses.
<sup>a</sup> The state enterprise office was included as a workplace. <sup>b</sup> Included restaurants, markets, malls, religious places, public places, and households of index patients or other people but not living together. <sup>c</sup> During the contact period. <sup>d</sup> Sharing multi-serving dishes using communal serving utensils to portion food individually was categorized as not sharing dishes. <sup>e</sup> Included sharing electronic cigarettes and any vaping devices. <sup>f</sup> Included washing with soap and water, and with alcohol-based solutions.

### Population attributable fraction

We estimated that the proportional reduction in cases that might occur if everyone wore a mask all the time during contact with COVID-19 patients (i.e., PAF of not wearing masks all the time) was 0.28 (Table 4). Among modifiable risk factors evaluated, PAF of shortest distance of contact <1 meter was highest at 0.40. If everyone wore a mask all the time; washed hands often; did not share a dish, cup, or cigarette; had shortest distance of contact >1 meter; and had duration of contact <15 min, cases may have been reduced by 84%, based on our data.

**Table 4.** Population attributable fraction (PAF) of risk factors for COVID-19 infection based on contact tracing data, Thailand, March-April 2020.

| Risk factors | Nightclub |  | Boxing stadium |  | Workplace |  | Household |  | Other places |  | Overall |  |
| --- | --- | --- | --- | --- | --- | --- | --- | --- | --- | --- | --- | --- |
|  | Prev<sup>a</sup> | PAF<sup>b</sup> | Prev<sup>a</sup> | PAF<sup>b</sup> | Prev<sup>a</sup> | PAF<sup>b</sup> | Prev<sup>b</sup> | PAF<sup>b</sup> | Prev<sup>a</sup> | PAF<sup>b</sup> | Prev<sup>a</sup> | PAF<sup>b</sup> |
| **Non-modifiable** |  |  |  |  |  |  |  |  |  |  |  |  |
| Female gender | 0.51 | 0.08 | 0.13 | 0.002 | 0.40 | 0.03 | 0.68 | 0.09 | 0.40 | 0.08 | 0.45 | 0.03 |
| Age group >15 years old | 1.00 | 0.32 | 0.98 | 0.05 | 0.99 | 0.37 | 0.82 | 0.26 | 0.96 | 0.37 | 0.95 | 0.15 |
| **Modifiable** |  |  |  |  |  |  |  |  |  |  |  |  |
| Distance of contact <1 m<sup>c</sup> | 0.88 | 0.71 | 0.98 | 0.19 | 0.65 | 0.72 | 0.87 | 0.68 | 0.85 | 0.76 | 0.82 | 0.40 |
| Duration of contact within 1 m >15 min<sup>c</sup> | 0.86 | 0.55 | 0.99 | 0.11 | 0.70 | 0.57 | 0.91 | 0.53 | 0.91 | 0.64 | 0.85 | 0.29 |
| Sharing dishes or cups<sup>c,d</sup> | 0.34 | 0.10 | 0.30 | 0.01 | 0.19 | 0.06 | 0.57 | 0.11 | 0.26 | 0.13 | 0.33 | 0.04 |
| Sharing cigarettes<sup>c,e</sup> | 0.08 | 0.12 | 0.02 | 0.001 | 0.01 | 0.06 | 0 | 0 | 0.01 | 0.007 | 0.02 | 0.02 |
| Not washing hands<sup>c,f</sup> | 0.05 | 0.06 | 0.21 | 0.01 | 0.20 | 0.17 | 0.10 | 0.08 | 0.28 | 0.29 | 0.16 | 0.04 |
| Not wearing masks all the time<sup>c,g</sup> | 0.60 | 0.52 | 0.80 | 0.08 | 0.78 | 0.65 | 0.86 | 0.55 | 0.94 | 0.68 | 0.78 | 0.28 |
| **Sum of all modifiable risk factors<sup>i</sup>** |  | 0.98 |  | 0.75 |  | 0.98 |  | 0.97 |  | 0.99 |  | 0.84 |

<sup>a</sup> Prevalence (Prev) was estimated using the imputed data set. <sup>b</sup> PAF was estimated using the direct method (Supplemental Text). <sup>c</sup> During the contact period. <sup>d</sup> Sharing multi-serving dishes using communal serving utensils to portion food individually was categorized as not sharing dishes. <sup>e</sup> Included sharing an electronic cigarette and any vaping device. <sup>f</sup> Washing hands included washing with soap and water, and with alcohol-based solutions. <sup>g</sup> Wearing masks incorrectly (i.e. not covering both nose and mouth) was considered to be the same as not wearing a mask for analyses. <sup>i</sup> Age and gender were considered non-modifiable risk factors, while other risk factors were considered modifiable. Total PAF was directly estimated using logistic regression in the form of natural logarithm; therefore, total PAF was not equal to the direct summation of PAF of each risk factor.

## DISCUSSION

Our findings provide evidence that mask-wearing, handwashing, and social distancing are independently associated with lower risk of COVID-19 infection among the general public in Thai community settings. We observed that wearing masks throughout the period of exposure to someone infected with COVID-19 was associated with lower risk of infection, while wearing masks sometimes during the exposure period was not. This evidence supports mask-wearing consistently and correctly at all times in public (*2, 7-9*).

We also quantified the effectiveness of different measures that could be implemented to prevent transmission in nightclubs, stadiums, workplaces, and other public gathering places. We estimated that adopting all recommendations (mask-wearing all the time; handwashing often; not sharing dishes, cups or cigarettes; maintaining a distance of >1 meter and, if this distance cannot be maintained, limiting contact duration to <15 minutes) might prevent up to 84% of COVID-19 infections in the study settings during the study period. Public gathering places may consider multiple measures to protect against COVID-19 and new pandemic diseases in the future.

The effectiveness of mask-wearing observed in this study is consistent with previous studies, including a randomized-controlled trial showing that consistent face mask use reduced risk of influenza-like illness (*28*); two case-control studies that found mask-wearing was associated with lower risk of SARS infection (*29, 30*); and a retrospective cohort study that found that wearing mask-wearing by index patients or family members at home was associated with lower risk of COVID-19 infection (*31*).

While previous studies found use of surgical masks or 12–16-layer cotton masks demonstrated protection against coronavirus infection in the community (*32*), we did not observe a difference between wearing non-medical and medical masks in the general population. Our results suggest that wearing non-medical masks in public can potentially reduce transmission of COVID-19 infection. Previous studies have shown perception of risk of developing COVID-19 can increase individuals’ likelihood of wearing medical masks in non-medical settings (*33*); however, given recent shortages, it is important that medical masks be reserved for use by healthcare workers.

We found a negative association between risk of COVID-19 infection and social distancing (i.e., shorter distances and durations of contact), consistent with previous studies that found that ≥1-meter physical distancing was associated with a larger protective effect, and distances of >2 meters could be even more effective (*32*). Our findings on effectiveness of hand hygiene were also consistent with those reported in previous studies (*34*).

Secondary attack rates at different venues in this study varied widely. The household secondary attack rate in our study (17%) is comparable with ranges reported previously (11%–23%) (*35, 36*) and relatively high compared to workplaces and other settings. While quarantine measures may be challenging and sometimes impractical, household members should immediately separate a person who develops any symptoms of COVID-19 from other household members (i.e., a sick person should stay in a specific room; use a separate bathroom, if possible; and not share dishes, cups, and other utensils) (*37*). All household members should wear masks, frequently wash hands, and perform social distancing to the extent possible (*38*).

The high number of COVID-19 patients associated with nightclub exposures in Bangkok is comparable to the COVID-19 outbreak associated with the Itaewon nightclub cluster in Seoul, Korea, in May 2020 (*39*). Similarly, we found individuals who visited several nightclubs in the same area during a short period of time. The secondary attack rate in boxing stadiums was high, at 86%. The high number of COVID-19 infections reported from a boxing stadium in Bangkok is similar to a cluster of COVID-19 cases associated with a football match in Italy in February 2020 (*40*). The secondary attack rate of COVID-19 at a choir practice in the United States was reported to be as high as 53% (*41*). Secondary attack rates in public gathering places with high densities of people who are shouting and cheering—such as football and boxing stadiums—are still uncertain but may be high.

It is likely that clear, consistent public messaging from policymakers can prevent a false sense of security and promote compliance with social distancing in Thailand. We found that those who wore masks throughout the exposure period to a case were also more likely to wash hands and perform social distancing. Both traditional and social media outlets can support public health responses by working with governments to provide consistent, simple and clear messages (*42*). In Thailand, daily briefings by Thailand’s Centre for COVID-19 Situation Administration (CCSA) provided clear and consistent messages on social distancing every day, including how to put on a mask and wash hands. CCSA’s regular situation reports and advice may have improved public confidence and compliance with the recommendations. Public messages on how to wear masks correctly need to be consistently delivered, particularly to those who wear masks sometimes or incorrectly (e.g., not covering both nose and mouth); we found that those who wear masks during exposure intermittently also did not practice social distancing adequately.

Our study has several limitations. First, our findings might not be generalizable to all settings, since they were based on contacts related to three major COVID-19 clusters in Thailand during March 2020. Second, estimated ORs were conditioned on occurrence of the reported contact with index patients. Our study did not evaluate or consider the probability of having contact with other infected people in the community setting, which must have occurred. Third, since only 89% of controls were tested at least once, those not tested could have been infected, and therefore, cases with mild or no symptoms who did not report symptoms or seek care and testing could have been missed. Fourth, it is impossible to identify every potential contact an individual has and some individuals may have had contact with >1 COVID-19 patient. Hence, our estimated secondary attack rates among contacts with high-risk exposure could be over- or underestimated. Fifth, findings were subject to common biases of retrospective case-control studies, including memory bias, observer bias, and information bias (*43*). To reduce potential biases, we used structured interviews where each participant was asked the same set of defined questions.

As social distancing measures are being relaxed in many countries, our findings provide evidence supporting consistent mask-wearing, handwashing, and adhering to social distancing recommendations can reduce COVID-19 transmission in public gathering places. Wearing non-medical masks in public may help slow the spread of COVID-19. The effectiveness of complying with all measures could be high; however, in places with a high density of people, additional measures may be required.

Clear and consistent public messaging on recommendations against COVID-19 infection is important, particularly targeting those who wear masks intermittently or incorrectly. Our data also showed that no single protective measure was associated with complete protection from COVID-19 infection. All measures, including wearing masks, handwashing, and social distancing can increase protection against COVID-19 infections in public.

## Data Availability

All data in aggregate are reported in the manuscript.

## Acknowledgements

We thank all participants and all COVID-19 patients involved in providing information. We thank all SRRT members at the central, regional, provincial and district levels, as well as all Village Health Volunteers in Thailand. We thank Pattraporn Klanjatturat for her technical assistance. We thank Dr. Suwannachai Wattanayingcharoenchai, Dr. Sombat Thanprasertkul, Dr. Virasakdi Chongsuvivatwong, Dr. Panithee Thammawijaya, and Dr. Walairat Chaifoo of the DDC, MoPH, and Dr. Virsasakdi Chongsuvivatwong from Prince of Songkhla University for their advice and direction.

## Contributors

PD, RS, CN, and DL contributed to design of the study. PD, RS, and DL contributed to setting up the database and quality control. AP, CJ, DR, ND, NE, NP, NS, OY, PaP, PiP, PK, PS, PW, SC, SK, and TC contributed to data collection. DL carried out the main statistical analysis. PD and RS coordinated the study and contributed to the statistical analyses. PD, RS, EB, and DL contributed to interpretation of the results and drafted the manuscript. All authors commented on drafts and read and approved the final manuscript. The corresponding author attests that all listed authors meet authorship criteria and that no others meeting the criteria have been omitted. PD is the guarantor.

## Competing interests

The authors declare that they have no completing interests.

## Ethical approval

As this study was part of the routine situation analysis and outbreak investigation of the DDC MoPH Thailand, it was not required to obtain ethics approval and no written informed consent was collected. However, the study team strictly followed ethical standards in research, that is, all individual information was strictly kept confidential and not reported in the paper. The DDC MoPH Thailand approved the analysis and reporting of data in aggregate.

## Funding

The study was supported by the DDC, MoPH, Thailand. DL is supported by the Wellcome Trust (106698/Z/14/Z).

## Data sharing

All data in aggregate are reported in the manuscript.

## Supplemental Materials

### Supplemental Methods

#### Study design

All high-risk contacts with any symptoms were tested with a reverse transcription polymerase chain reaction (RT-PCR) assay and quarantined in a hospital or a quarantine site. All high-risk contacts without any symptoms were self-quarantined at home. Before 23 March 2020, all high-risk contacts without any symptoms were tested using RT-PCR assays on day 5 after the last date of exposure to a case (*19, 20*). As of 23 March 2020, all household contacts were tested using RT-PCR assays regardless of their symptoms (*19, 20*). Other high-risk contacts were tested only if they developed any COVID-19 symptoms. All low-risk contacts were recommended to perform self-monitoring for 14 days, and visit healthcare facilities immediately for RT-PCR assays if they developed any symptoms of COVID-19 (*19, 20*).

**STROBE Statement**Checklist of items that should be included in reports of ***case-control studies***

|  | Item No | Recommendation | Page No |
| --- | --- | --- | --- |
| **Title and abstract** | 1 | (a) Indicate the study's design with a commonly used term in the title or the abstract | 1-3 |
|  |  | (b) Provide in the abstract an informative and balanced summary of what was done and what was found | 1-3 |
| **Introduction** |  |  |  |
| Background/rationale | 2 | Explain the scientific background and rationale for the investigation being reported | 4-5 |
| Objectives | 3 | State specific objectives, including any prespecified hypotheses | 4-5 |
| **Methods** |  |  |  |
| Study design | 4 | Present key elements of study design early in the paper | 6-9 |
| Setting | 5 | Describe the setting, locations, and relevant dates, including periods of recruitment, exposure, follow-up, and data collection | 6-9 |
| Participants | 6 | (a) Give the eligibility criteria, and the sources and methods of case ascertainment and control selection. Give the rationale for the choice of cases and controls | 6-9 |
|  |  | (b) For matched studies, give matching criteria and the number of controls per case | NA |
| Variables | 7 | Clearly define all outcomes, exposures, predictors, potential confounders, and effect modifiers. Give diagnostic criteria, if applicable | 6-9 |
| Data sources/<br>measurement | 8* | For each variable of interest, give sources of data and details of methods of assessment (measurement). Describe comparability of assessment methods if there is more than one group | 6-9 |
| Bias | 9 | Describe any efforts to address potential sources of bias | 6-9 |
| Study size | 10 | Explain how the study size was arrived at | 6-9 |
| Quantitative variables | 11 | Explain how quantitative variables were handled in the analyses. If applicable, describe which groupings were chosen and why | 6-9 |
| Statistical methods | 12 | (a) Describe all statistical methods, including those used to control for confounding | 6-9 |
|  |  | (b) Describe any methods used to examine subgroups and interactions | 9 |
|  |  | (c) Explain how missing data were addressed | 6-9 |
|  |  | (d) If applicable, explain how matching of cases and controls was addressed | NA |
|  |  | (e) Describe any sensitivity analyses | 9 |
| **Results** |  |  |  |
| Participants | 13* | (a) Report numbers of individuals at each stage of study—eg numbers potentially eligible, examined for eligibility, confirmed eligible, included in the study, completing follow-up, and analysed | 10-12 |
|  |  | (b) Give reasons for non-participation at each stage | 10-12 |
|  |  | (c) Consider use of a flow diagram | Figure1 |
| Descriptive data | 14* | (a) Give characteristics of study participants (eg demographic, clinical, social) and information on exposures and potential confounders | 10-12<br>Table2 |
|  |  | (b) Indicate number of participants with missing data for each variable of interest | Table2 |
| Outcome data | 15* | Report numbers in each exposure category, or summary measures of exposure | 10-12 |
| Main results | 16 | (a) Give unadjusted estimates and, if applicable, confounder-adjusted estimates and their precision (eg, 95% confidence interval). Make clear which confounders were adjusted for and why they were included | Table2 |
|  |  | (b) Report category boundaries when continuous variables were categorized | 10-12<br>Table2 |
|  |  | (c) If relevant, consider translating estimates of relative risk into absolute risk for a meaningful time period |  |
| Other analyses | 17 | Report other analyses done—eg analyses of subgroups and interactions, and sensitivity analyses | 12-13<br>Table4 |
| **Discussion** |  |  |  |
| Key results | 18 | Summarise key results with reference to study objectives | 13 |
| Limitations | 19 | Discuss limitations of the study, taking into account sources of potential bias or imprecision. Discuss both direction and magnitude of any potential bias | 16 |
| Interpretation | 20 | Give a cautious overall interpretation of results considering objectives, limitations, multiplicity of analyses, results from similar studies, and other relevant evidence | 14-17 |
| Generalisability | 21 | Discuss the generalisability (external validity) of the study results | 14-17 |
| **Other information** |  |  |  |
| Funding | 22 | Give the source of funding and the role of the funders for the present study and, if applicable, for the original study on which the present article is based | 18 |

\*Give information separately for cases and controls.
**Note:** An Explanation and Elaboration article discusses each checklist item and gives methodological background and published examples of transparent reporting. The STROBE checklist is best used in conjunction with this article (freely available on the Web sites of PLoS Medicine at http://www.plosmedicine.org/, Annals of Internal Medicine at http://www.annals.org/, and Epidemiology at http://www.epidem.com/). Information on the STROBE Initiative is available at http://www.strobe-statement.org.

#### Statistical analysis

We estimated the direct population attributable fraction (PAF) using the imputed dataset and the direct method as previously described (*26, 27*). Direct PAF can be obtained by calculating PAFs directly from individuals’ data using logistic regression (*26, 27*). First, we modified our final logistic regression model by considering each risk factor dichotomously. Then, irrespective of exposure to each risk factor for each individual, that factor was removed from the population by calculating probability based on all observations as unexposed. The predicted probability of developing COVID-19 infection for each asymptomatic contact, with the assumption that there was no exposure to a certain risk factor, was defined by:

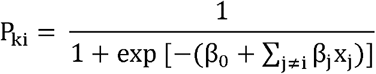

P_ki_ is representative of predicted probability of COVID-19 infection in individual asymptomatic contact k, assuming no exposure to a specific risk factor (x_i_); β_j_ indicates the regression coefficient of risk factor (x_j_), except risk factor number i (x_i_). Subsequently, the sum of all predicted probabilities for all individuals in the study would be equal to adjusted estimate of total cases, which is anticipated in the absence of that specific risk factor (x_i_).

Then, PAF was estimated by subtraction of the total number of predicted cases from total number of observed cases, divided by the total number of observed cases:

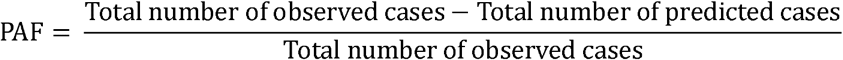

## Supplemental Results

For the nightclub cluster, we identified 11 primary index patients who started having symptoms from 4 to 8 March and were diagnosed (and isolated) from 3 to 10 March (Supplemental Figure 1). Those primary index patients visited multiple nightclubs included in the analysis during the study period, and 35 of 228 (15%) asymptomatic contacts at nightclubs had PCR-confirmed COVID-19 infections after the contact (Figure 2, Cluster A).

For the boxing stadium cluster, we identified 5 primary index patients who started having symptoms from 6 through 12 March and were diagnosed (and isolated) from 11 through 21 March (Supplemental Figure 2). Those primary index patients visited multiple boxing stadiums included in the analysis during the study period, and 125 of 144 (87%) asymptomatic contacts at the boxing stadiums had RT-PCR-confirmed COVID-19 infections after the contact (Figure 2, Cluster B).

Among the two primary index patients for the office cluster, one had had symptoms since 15 March 2020 (Primary index patient C1 in Supplemental Figure 3) and was considered the source of infection for one new case in the office during the study period. The other primary index patient (Primary index patient C2 in Supplemental Figure 3) was a household member of a staffer at the office and was considered as the source of infection for the staffer via household contact.

**Supplemental Table 1.** **Factors associated with COVID-19 infection in a multivariable model including type of mask among persons followed through contract tracing, Thailand, March-April 2020**

| Factors | Adjusted odds ratio (95% CI)<sup>a</sup> | P |
| --- | --- | --- |
| **Male gender** | 0.75 (0.40–1.38) | 0.35 |
| **Age group** |  |  |
| ≤15 years old | 0.56 (0.14–2.18) | 0.19 |
| >15–40 years old | 1.0 |  |
| >40–65 years old | 1.77 (0.94–3.35) |  |
| >65 years old | 1.00 (0.23–4.34) |  |
| **Contact place<sup>b</sup>** |  |  |
| Nightclub | Not applicable<sup>c</sup> | - |
| Boxing stadium |  |  |
| Workplace |  |  |
| Household |  |  |
| Others |  |  |
| **Shortest distance of contact** |  |  |
| Physical contact | 1.0 | 0.02 |
| ≤1 meter without physical contact | 1.08 (0.57–2.03) |  |
| >1 meter | 0.15 (0.04–0.66) |  |
| **Duration of contact within 1 meter** |  |  |
| >60 minutes | 1.0 | 0.10 |
| >15–60 minutes | 0.67 (0.29–1.56) |  |
| ≤15 minutes | 0.25 (0.07–0.94) |  |
| **Sharing dishes or a cups<sup>d,e</sup>** |  |  |
| None | 1.0 | 0.39 |
| Yes | 1.32 (0.69–2.53) |  |
| **Sharing cigarettes<sup>d,f</sup>** |  |  |
| None | 1.0 | 0.03 |
| Yes | 3.48 (1.09–11.05) |  |
| **Washing hands<sup>d,g</sup>** |  |  |
| None | 1.0 | 0.04 |
| Sometimes | 0.33 (0.14–0.79) |  |
| Often | 0.33 (0.12–0.87) |  |
| **Wearing masks<sup>d,h</sup>** |  |  |
| Not wearing masks | 1.0 | 0.54 |
| Wearing non-medical masks | 1.29 (0.48–3.45) |  |
| Wearing non-medical and medical masks alternately | 1.03 (0.26–4.07) |  |
| Wearing medical masks | 0.61 (0.25–1.49) |  |
| **Wearing masks all the time<sup>d,h</sup>** |  |  |
| No | 1.0 | 0.02 |
| Yes | 0.32 (0.12–0.82) |  |

<sup>a</sup> Both crude and adjusted odds ratios were estimated using logistic regression with a random effect for location and a random effect for index patient nested within the same location. Missing values were imputed using the imputation model. <sup>b</sup> The state enterprise office was included in workplaces. Others included restaurants, markets, malls, religious places, and households of index patients or other people but not living together, etc. <sup>c</sup> Location was included in the model as a random effect variable. <sup>d</sup> During the contact period. <sup>e</sup> Sharing multi-serving dishes using communal serving utensils to portion food individually was categorized as not sharing dishes. <sup>f</sup> Included sharing electronic cigarettes and any vaping devices. <sup>g</sup> Included washing with soap and water, and with alcohol-based solutions. <sup>h</sup> Wearing masks incorrectly (e.g., not covering both nose and mouth) was considered to be the same as not wearing a mask for analyses.

**Supplemental Figure 1.**
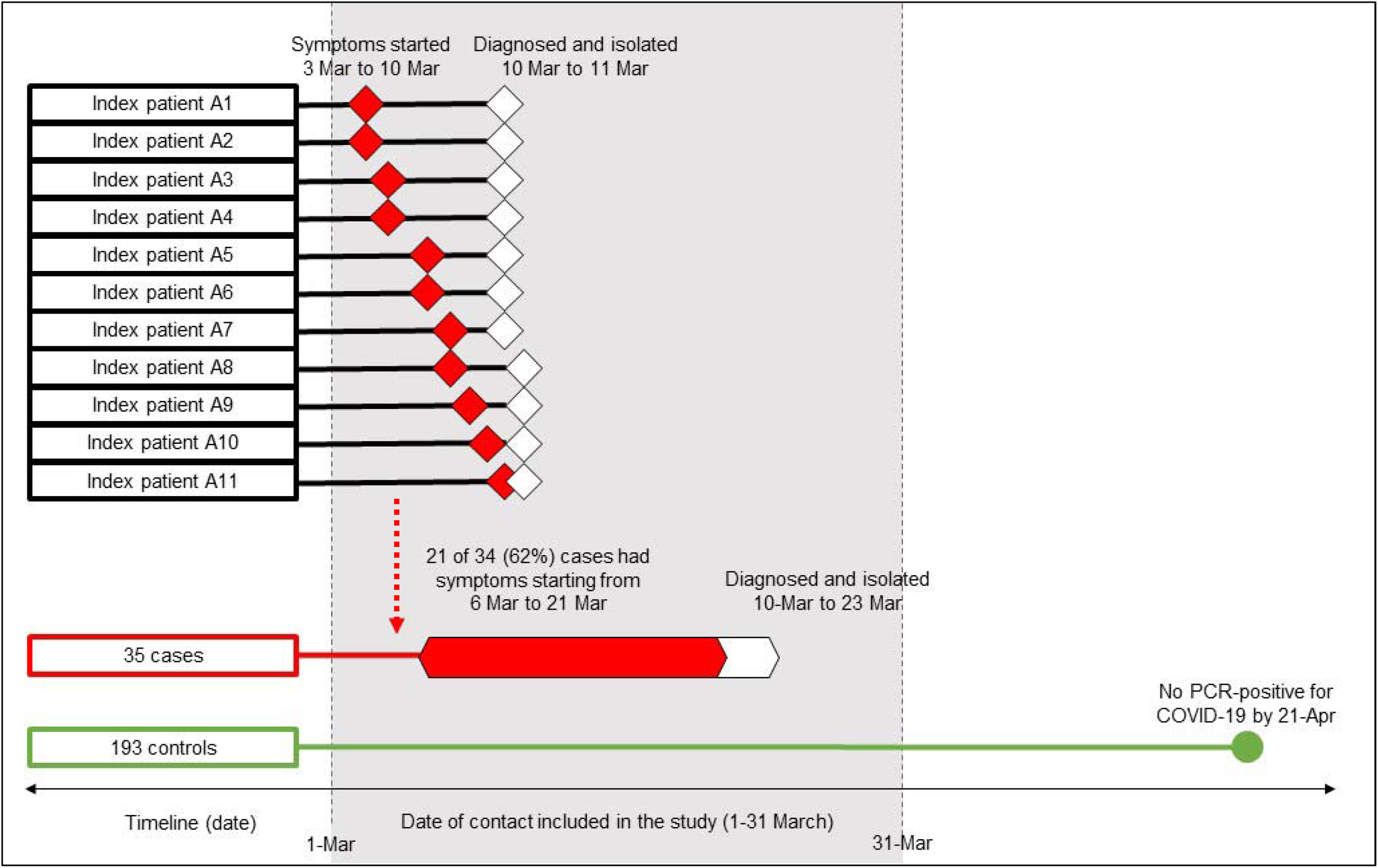
**Timeline and possible SARS-CoV2 transmission of primary index patients of the nightclub cluster Thailand, March-April 2020**

**Supplemental Figure 2.**
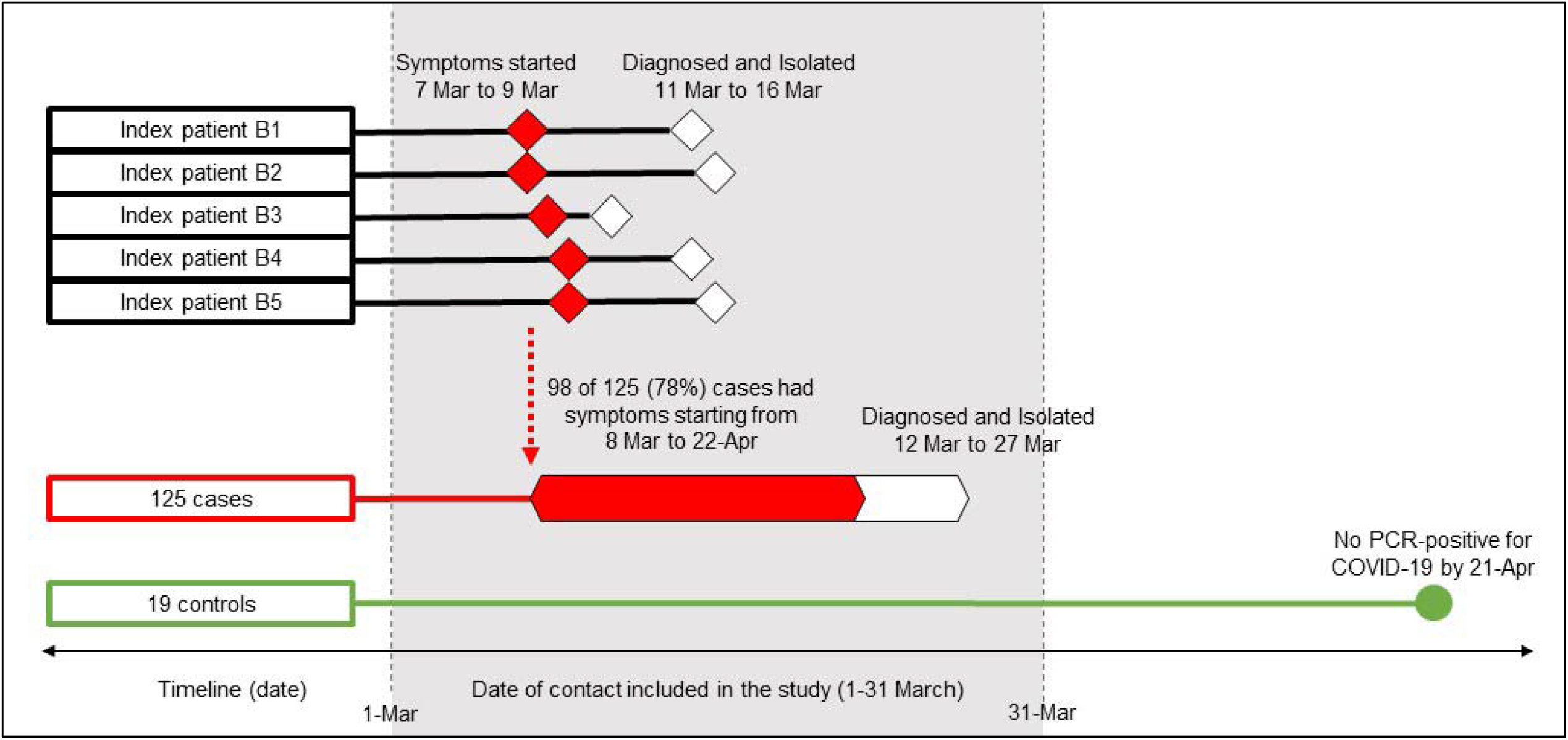
**Timeline and possible SARS-CoV2 transmission of primary index patients of the boxing stadium cluster, Thailand, March-April 2020**

**Supplemental Figure 3.**
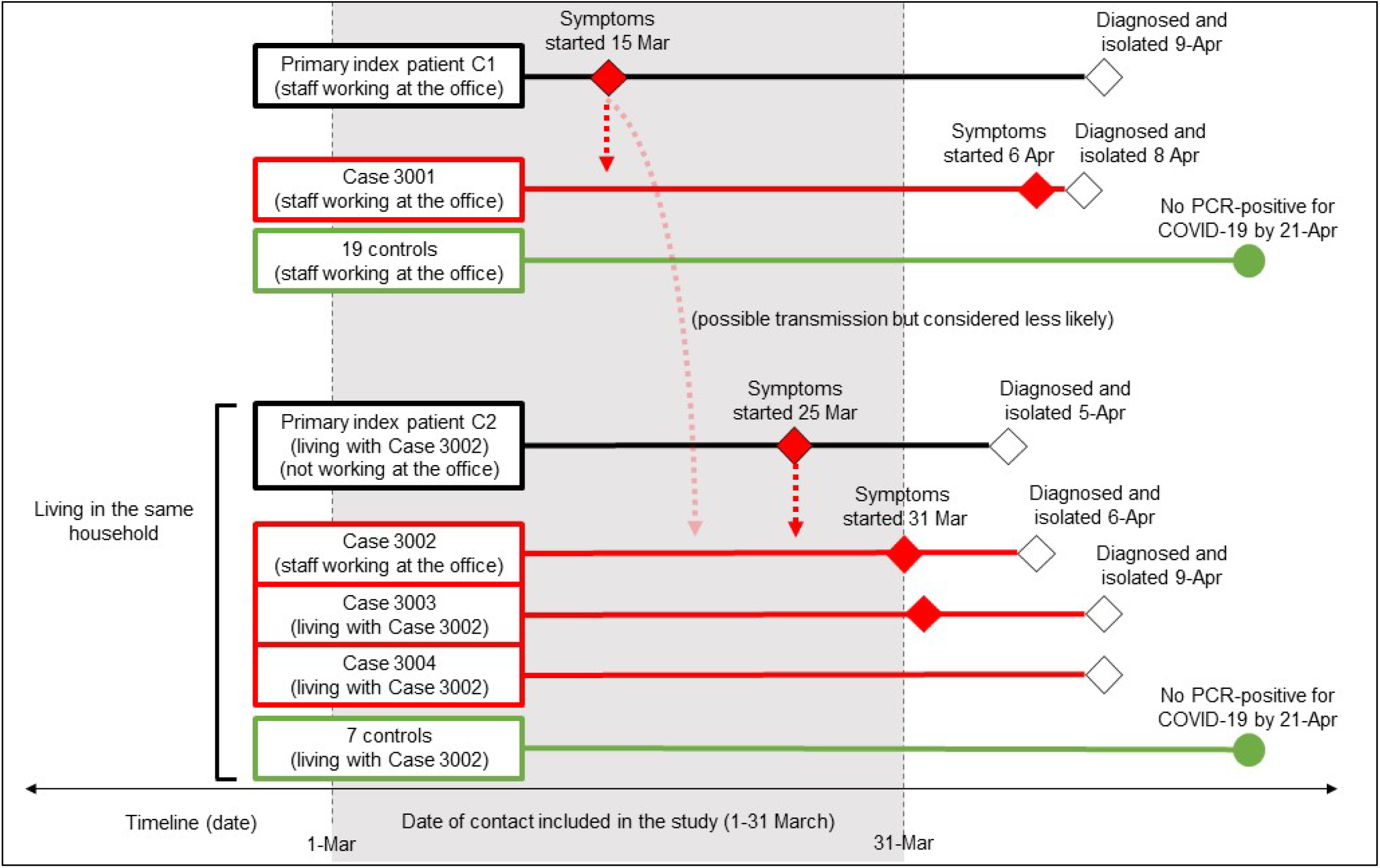
**Timeline and possible SARS-CoV2 transmission of primary index patients of the state enterprise office cluster, Thailand, March-April 2020**

## Notes

### Competing Interest Statement

The authors have declared no competing interest.

### Funding Statement

The study was supported by the Department of Disease Control (DDC), Ministry of PUblic Health (MoPH) Thailand. DL is supported by the Wellcome Trust (106698/Z/14/Z).

### Summary of Updates

The revised manuscript has received clearance from Centers for Disease Control and Prevention headquarter, and an author (Dr Emily Bloss from Thailand Ministry of Public Health - U.S. Centers for Disease Control and Prevention Collaboration) has been included. All co-authors approved the revised manuscript and the addition of Dr Emily Bloss as an author. There are no major changes in study design, analyses and presentations.

